# Affected medical services in Iwate prefecture in the absence of a COVID-19 outbreak

**DOI:** 10.1101/2020.06.19.20135269

**Authors:** Noriyuki Sasaki, Satoshi S. Nishizuka

## Abstract

The Japanese government has declared a national emergency and travel entry ban since the coronavirus disease 2019 (COVID-19) pandemic began. As of June 19, 2020, there have been no confirmed cases of COVID-19 in Iwate. Here, we analyzed the excess deaths as well as the number of patients and medical earnings due to the pandemic from prefectural hospitals located in one of the least-affected areas in Japan. From January to March 2020, the excess death rates per month were not significantly higher than the past-year average. Compared to January 2020, the numbers of both outpatients and inpatients in April 2020 showed a 5.2% and 6.1% decrease, respectively. Accordingly, the amount of medical earnings of both outpatients and inpatients in April 2020 showed a 3.0% and 6.3% decrease, respectively. Present analysis demonstrated that there were no excess deaths due to “unidentified” COVID-19 infections in Iwate; however, hospital budgetary management has been affected by the social restrictions. Regardless of COVID-19 infection spread, it may be difficult to maintain daily medical services if such low service activity continues in the existing hospitals. Additional longitudinal studies will be necessary to evaluate the effects of social restrictions on hospital management, but the true demand of regional medical services may emerge after this outbreak.

## Introduction

Iwate prefecture is one of the Japanese municipal governments located along the northeastern Pacific coast, where approximately 1.26 million people live in an area equivalent to the state of Connecticut. In Iwate, there are 1,376 beds *per capita* and 26.7% have been maintained by 20 prefectural hospitals in 2018^1^. As of June 19, 2020, there have been no confirmed cases of coronavirus disease 2019 (COVID-19) in Iwate.^2^ The nationwide excess death rate during the pandemic has been 0.7% below the historical average.^3^ These results suggest that immediate massive disruption of medical care by the COVID-19 outbreak is unlikely to occur in Iwate.

The Japanese government has declared a national emergency and travel entry ban for >180 countries/regions since the pandemic began. These restrictions have resulted in suppression of travel, logistics, and consumer confidence. Subsequently, the financial impact on some businesses has been seen despite a decline in the number of COVID-19 infections. It can now be hypothesized that even public hospitals may face a financial burden. Here, we analyzed the excess deaths as well as the number of patients and medical earnings due to the pandemic from prefectural hospitals located in one of the least-affected areas of municipal governments in Japan.

## Methods

The data analytical period was between February 2019 and April 2020. The number of deaths in Iwate was obtained from a public database released by the Japanese Ministry of Health, Labor and Welfare. The excess death rate in Iwate was defined as follows: the number of deaths per month in 2020 divided by the past-year (2015-2019) average number of deaths per month. The number of patients and medical earnings in all 20 Iwate prefectural hospitals and 6 clinics was obtained with permission from the Iwate Prefecture Medical Bureau. A three-month moving average was calculated in terms of the number of outpatients and inpatients as well as the medical earnings of outpatients and inpatients.

## Results

From January to March 2020, the excess death rates per month in Iwate were not significantly higher than the past-year average (Fig. 1A), whereas the numbers of patients and amount of medical earnings per month showed a decreasing trend (Fig. 1B, C). Compared to January 2020, the numbers of both outpatients and inpatients in April 2020showed a 5.2% and 6.1% decrease, respectively (Fig. 1B). Accordingly, the amount of medical earnings of both outpatients and inpatients in April 2020 showed a 3.0% and 6.3% decrease, respectively (Fig. 1C). All major governmental social declarations were released between January and April 2020.

**Figure.**
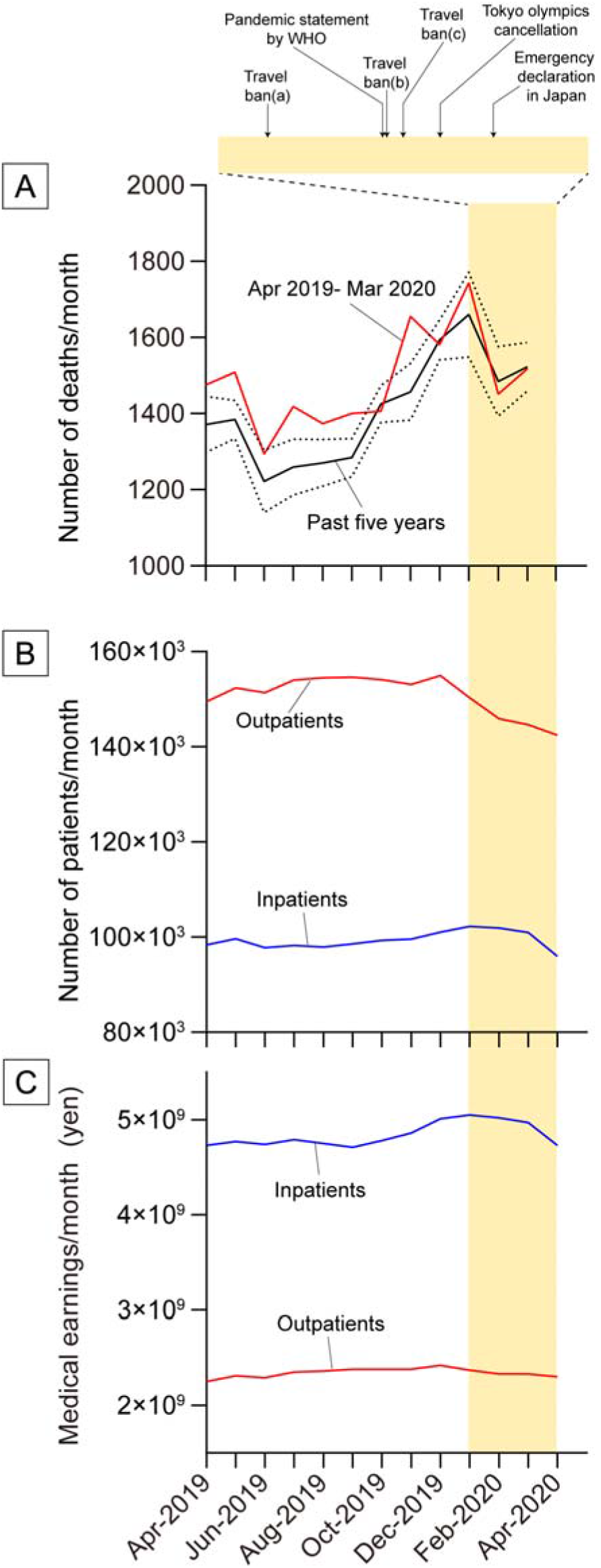
Longitudinal monitoring parameters representing deaths and patients in prefectural hospitals in Iwate. (A) Monthly monitoring of excess deaths. The dotted lines show the 95% confidence intervals of the average in the past five years. Governmental or public statements made during the outbreak are listed on top. Travel ban warnings were released for China (a), United States (b) and other countries (c). (B) The total number of outpatients and inpatients between April 2019 and April 2020. (C) The total amount of medical earnings from April 2019 to April 2020.

## Discussion

Unlike a city lockdown, Japan’s emergency declaration is not intended to be an extraordinary compulsory regulation. Present analysis demonstrated that there were no excess deaths due to “unidentified” COVID-19 infections in Iwate; however, hospital budgetary management has been affected by the emergency declaration. Routine hospital visits were postponed for outpatients and non-urgent surgeries were rescheduled. A hospital visit itself is considered to be a risk for infection. Regardless of COVID-19 infection spread, it may be very difficult to maintain daily medical services if such low service activity continues in the existing hospitals. Limitations of this study include a limited follow-up period through April 30, 2020 and the lack of appropriate municipal governments in Japan for comparison. In addition, the excess deaths were counted monthly instead of weekly because of public data availability^4^. Thus, the present data may not be fully applicable to specific issues in individual hospitals.

Additional longitudinal studies will be necessary to evaluate the effects of social restrictions on hospital management, but the true demand of regional medical services may emerge after this outbreak.

## Data Availability

Consider upon request.

## Article Information

### Author Contributions

Concept and design: Nishizuka.

Acquisition, analysis, or interpretation of data: Sasaki

Drafting of the manuscript: Sasaki and Nishizuka.

Statistical analysis: Sasaki.

Supervision: Nishizuka.

### Conflict of Interest Disclosures

Nishizuka received grant/research support form Taiho Pharmaceuticals,

Boehringer-Ingelheim, Chugai Pharmaceutical, and Iwate prefecture; and is a part-time

employee of Iwate Prefectural Hospitals. No other disclosures were reported.

### Additional Contributions

We thank Manabu Ogasawara of Iwate Medical Bureau for the data acquisition.

